# A Qualitative Study Exploring the Consumer Experience of Receiving Self-Initiated Polygenic Risk Scores from a Third-Party Website

**DOI:** 10.1101/2022.07.04.22277219

**Authors:** Kiara Lowes, Kennedy Borle, Lasse Folkersen, Jehannine Austin

**Author notes:** Corresponding author: Jehannine Austin, MSc, PhD, A3-127 939 W28th Ave, Vancouver, British Columbia, Canada.

## Abstract

The number of people accessing their own polygenic risk scores (PRSs) online is rapidly increasing, yet little is known about why people are doing this, how they react to the information, and what they do with it. We conducted a qualitative interview-based study with people who pursued PRSs through Impute.me, to explore their motivations for seeking PRS information, their emotional reactions, and actions taken in response to their results. Using interpretive description, we developed a theoretical model describing the experience of receiving PRSs in a direct-to-consumer (DTC) context. Dissatisfaction with healthcare was an important motivator for seeking PRS information. Participants described having medical concerns dismissed, and experiencing medical distrust, which drove them to self-advocate for their health, which in turn ultimately led them to seek PRSs. Polygenic risk scores were often empowering for participants, but could be distressing when PRS information did not align with participants’ perceptions of their personal or family histories. Behavioural changes made in response to PRS results included dietary modifications, changes in vitamin supplementation and talk-based therapy. Our data provides the first qualitative insight into how people’s lived experience influence their interactions with DTC PRSs.

## INTRODUCTION

Polygenic risk scores (PRSs) are an emerging technology that have ignited debates in the field of genetics about the possibilities and limitations of genetic testing for complex conditions(1–5). Polygenic risk scores represent a sum of weighted genetic risk factors, providing an estimate of an individual’s genetic predisposition for a complex condition. While PRSs are not yet being provided in clinical settings, there are moves towards the integration of PRSs into clinical care to guide screening and prevention strategies(6). Direct-to-consumer (DTC) PRSs have also become available via third party websites that allow people to upload their genetic information to obtain PRS information(7). Despite the rapid uptake of DTC PRSs, limited research has examined the issues and opportunities related to seeking PRSs in this context.

Studies exploring the impacts of other (non-PRS) types of DTC genetic testing (DTC-GT) have reported either no change in health-related distress levels(8–11) or a reduction in anxiety(12). Importantly, many of these studies involved genetic counseling or the provision of *hypothetical* genetic test results, making them less representative of how consumers are accessing their genetic information in reality. Adverse reactions and decisional regret have been reported among people who have self-initiated non-PRS related DTC-GT(13–15). To date, PRS research has mainly focused on contexts where PRSs are provided to individuals with a personal or family history of cancer(16–21) or bipolar disorder(22). When provided in these settings, PRSs have served as an explanation for people’s personal or family history and reduce feelings of self-blame. When provided for conditions that are not tailored towards a family history, PRSs have not been found to elicit high levels of distress, though PRSs in this study were provided through a research protocol with informed consent(23). To our knowledge, only one study has examined the issues related to receiving *self-initiated* DTC PRSs where there is no healthcare provider involvement, finding that >50% of users experienced some degree of negative reaction, with 5% showing signs suggestive of post-traumatic stress disorder related to receiving their result(24).

We sought to develop a deeper understanding of the motivations for self-initiating PRS testing and reactions to results through a qualitative, interview-based study.

## MATERIALS AND METHODS

We conducted a qualitative study in which we interviewed self-initiators of PRSs to gain insight into their experience with seeking and receiving PRSs. An interpretive description approach was used for study design and data analysis. Interviews were conducted and coded by the first author (KL), who was a genetic counseling student at the time, and who identified as female. KL had no prior relationships with any of the research participants. Data interpretation was completed by KL, KB, and JA, who are all genetic counselors who do not provide PRSs clinically. None of the members of the study team who were involved in the collection or analysis of the data had personal experience with receiving personal genetic information via Impute.me. Written and verbal consent were obtained prior to all data collection. This study was approved by the University of British Columbia’s Research Ethics Board (H21-01616).

### Eligibility

Study participants were recruited from Impute.me, a not-for-profit website (https://impute.me) where users could upload their raw genetic information to obtain PRSs. Participants were eligible if they were (1) at least 19 years of age; (2) fluent in written and spoken English; and (3) had received one or more PRSs through the complex disease module of the website.

An email was distributed to all Impute.me users, providing a brief description of the research study followed by a short quantitative questionnaire including demographics, immediate response to PRS results, time elapsed since results were received, and present-day emotional impact of PRS information as measured by the Impact of Event Scale—Revised (IES-R)(25). Respondents were asked to provide their contact information if they wanted to be considered for an interview.

### Sampling approach

We used respondents’ questionnaires for purposive sampling to ensure broad representation of experiences (e.g. a range of IES-R scale score) and voices (e.g. participants from diverse racial or gender groups) in our interviews.

### Interviews

One-on-one, semi-structured interviews were conducted via Zoom by the first author. The interview guide (see supplemental material) was developed based on quantitative research previously conducted by the study team(24) to explore motivations for seeking PRSs, and emotional responses and actions taken after receiving PRS results. Interviews were audio-recorded and transcribed verbatim by the first author.

### Data analysis

Interview data was analyzed using interpretive description(26,27), which is a qualitative methodology that builds upon the practice of thematic analysis. Interpretive description analyzes the individual and collective experience of a study population to develop common themes that are assembled into a complex picture. The aim of this methodology is to develop a deep understanding of a human experience so as to inform clinical practice(26).

Throughout data collection, regular meetings were held to discuss concepts drawn from the interviews and to revise the interview guide to explore themes more thouroughly. Interview transcripts were coded and catalogued by the first author and reviewed with the study team. Several meetings were held to brainstorm relationships between themes and to build a theoretical model demonstrating the process of receiving DTC PRSs. The model was reviewed with three interview participants, who were asked to provide feedback on whether the model resonated with their experiences. Small changes were made to the model to reflect participant feedback.

## RESULTS

### Participants

Of the 56,280 people who were contacted about the study, 209 completed the survey with their contact information indicating their interest in an interview. We invited 23 participants to complete an interview and ultimately conducted 11 interviews, ranging from 21 to 69 minutes in length. Interviewee demographics are summarized in Table 1.

**Table 1.**
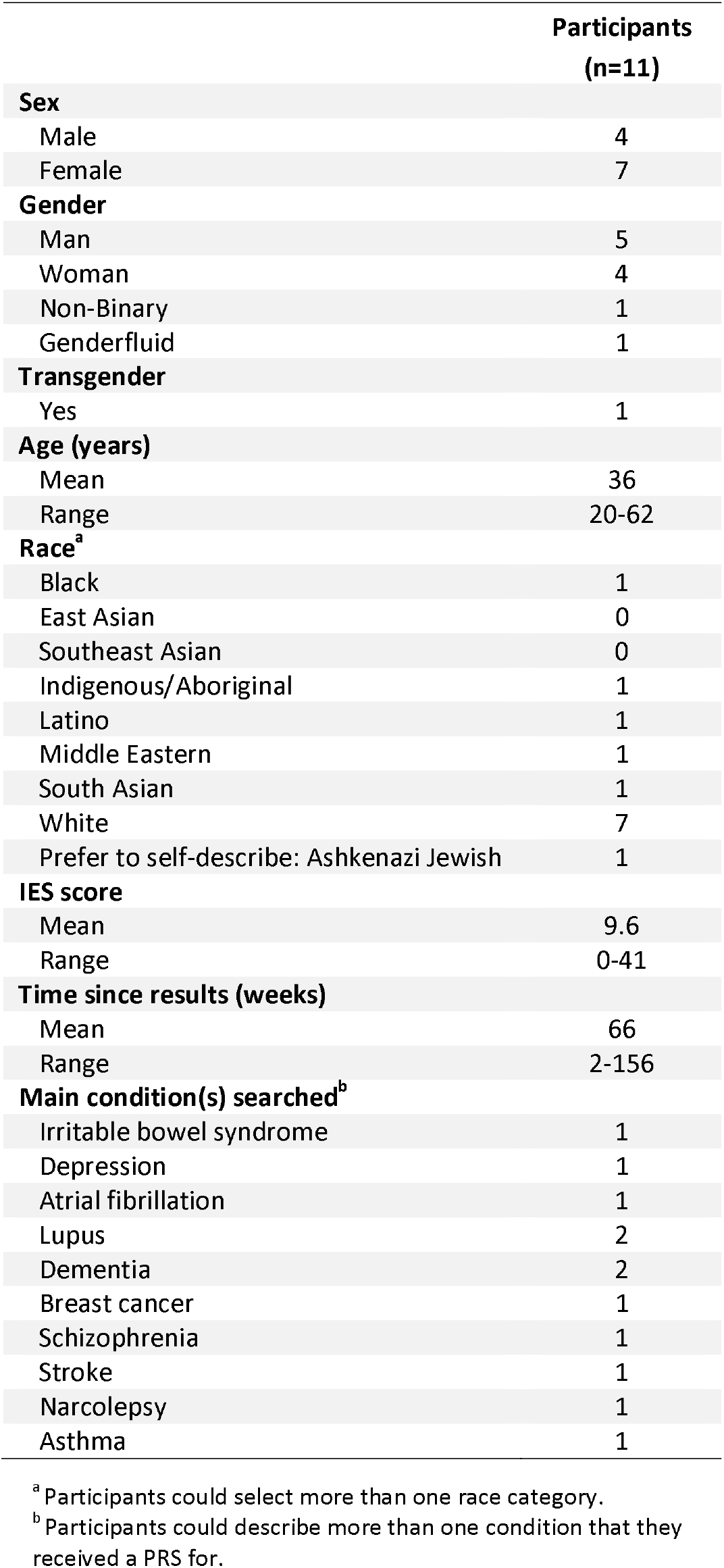
Demographics of interview participants.

|  | Participants<br>(n=11) |
| --- | --- |
| **Sex** |  |
| Male | 4 |
| Female | 7 |
| **Gender** |  |
| Man | 5 |
| Woman | 4 |
| Non-Binary | 1 |
| Genderfluid | 1 |
| **Transgender** |  |
| Yes | 1 |
| **Age (years)** |  |
| Mean | 36 |
| Range | 20-62 |
| **Race<sup>a</sup>** |  |
| Black | 1 |
| East Asian | 0 |
| Southeast Asian | 0 |
| Indigenous/Aboriginal | 1 |
| Latino | 1 |
| Middle Eastern | 1 |
| South Asian | 1 |
| White | 7 |
| Prefer to self-describe: Ashkenazi Jewish | 1 |
| **IES score** |  |
| Mean | 9.6 |
| Range | 0-41 |
| **Time since results (weeks)** |  |
| Mean | 66 |
| Range | 2-156 |
| **Main condition(s) searched<sup>b</sup>** |  |
| Irritable bowel syndrome | 1 |
| Depression | 1 |
| Atrial fibrillation | 1 |
| Lupus | 2 |
| Dementia | 2 |
| Breast cancer | 1 |
| Schizophrenia | 1 |
| Stroke | 1 |
| Narcolepsy | 1 |
| Asthma | 1 |

<sup>a</sup> Participants could select more than one race category.
<sup>b</sup> Participants could describe more than one condition that they received a PRS for.

### Overview of the theoretical model

Figure 1 shows a visual depiction of the theoretical model that was constructed through the study. A core driver for seeking PRS information was dissatisfaction with health care. Specifically, participants expressed that their medical concerns had been ignored (*dismissed medical concerns*) and/or felt that they had been inadequately treated by their healthcare providers (*medical distrust*). These experiences resulted in a medical void (*represented in Figure 1 by the unfilled icon representing a person*) that participants sought to fill through self-advocating for their own health, which led them to seek PRSs. Polygenic risk score information was often validating for participants, though for others, unexpected results led to shock and test-related distress. Many participants reported making behavioural changes, regardless of their reactions to their results.

**Figure 1.**
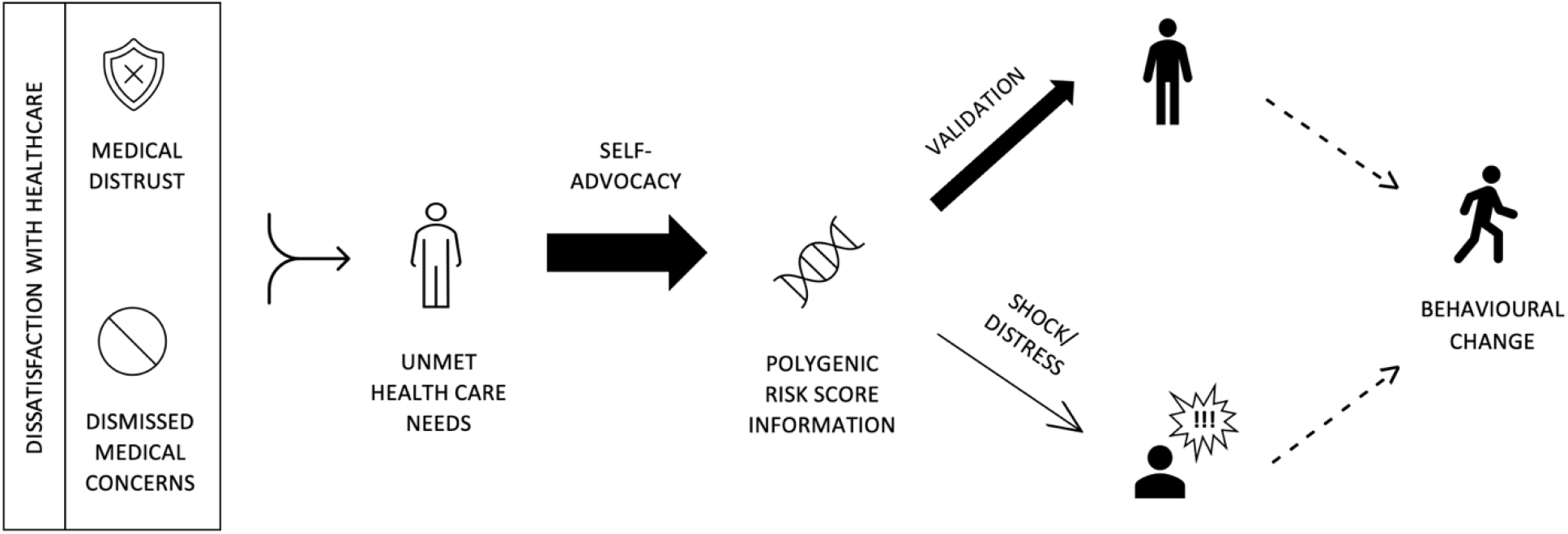
A visual representation of the process of obtaining polygenic risk scores (PRSs), beginning with the motivations for pursuing PRS information, how participants reacted to their results, and what they did with their results. Various arrow weights are used to demonstrate major and minor pathways taken by participants; thick arrows represent common routes and thin arrows represent less common routes. Dashed arrows are used to indicate routes that not everyone takes; this process may end with an emotional response, or some may proceed to make behavioural changes in response to their PRS results.

### Dissatisfaction with healthcare

A main factor that motivated people to seek PRS information was their dissatisfaction with the healthcare that they had received. This dissatisfaction was comprised of two components, medical distrust and dismissed medical concerns.

### Medical distrust

Many participants had experienced illness and felt like their treatment, or lack thereof, had only exacerbated their symptoms, which in turn created a sense of distrust in the medical system.

> *“They had me filled with all sorts of medications for high blood pressure and diabetes and other things, all those things just made me sicker*.*” – Participant 7*

### Dismissed medical concerns

Participants had experienced health concerns but felt dismissed when they brought these medical concerns to a healthcare provider – this led to dissatisfaction with their healthcare.

> *“Every time I felt sick I would go to a doctor and they would tell me there’s nothing wrong with me*.*” – Participant 4*

Several participants described a diagnostic odyssey which had not resulted in a satisfactory explanation for their illness.

> *“I think family doctors aren’t really great for this sort of stuff. They have a saying about how if you hear hooves think horses not zebras*… *But then, the problem is what if you have an unusual condition? Because the next step is to be dismissive, [say] it’s all in your head, or we don’t know what it is*.*” – Participant 7*

### Self-advocacy

Dissatisfaction with healthcare and unmet healthcare needs left a medical void, that often motivated participants to attempt to fulfill their own needs.

> *“I kept looking for answers instead of screaming for help… I screamed for help for months and no one came*.*” – Participant 4*

As self-described “information seekers,” participants advocated for themselves by seeking out genetic information.

> *“I’ve always been a generally curious person. So, if it was something that I could potentially figure out about myself that I couldn’t otherwise figure out, that’s enough for me to be curious about what I could find. Just not knowing would be too hard*.*” – Participant 11*

### Polygenic risk score information

There were varied reasons for why participants looked at specific types of PRSs. Some looked up PRSs for their own diagnoses – many did this for mental health conditions in particular. Participant 5 described their motivation for pursuing a PRS for their mental health condition: *“I wanted to confirm what was already there…it brought me closure*.*”*

Other people targeted their search to conditions that they exhibited symptoms of but did not have a formal diagnosis for, or they sought out PRSs for conditions that were running in their family. Others described looking up PRSs for everything that was offered on Impute.me, regardless of their personal or family history.

### Validation

Overall, participants were satisfied with their PRS results. Some people felt like this information allowed them to better understand themselves and their health.

> *“It’s like getting myself back… I understand the body I inhabit*.*” – Participant 3*

When participants received results that were consistent with their health history, they felt justified in their concerns.

> *“When people are dismissive of these things, it’s useful to have something that actually backs you up*.*” – Participant 7*

Participants who had concerns about their family history, found it helpful to have confirmation of their genetic predisposition. Participant 10 described how their PRS reduced the uncertainty surrounding their grandparent’s Alzheimer’s diagnoses:

> *“[It’s useful] knowing conclusively that I seem to have a higher genetic risk and that it’s not just whatever was in the ground water when my grandparents were growing up*.*”*

### Shock/Distress

When participants received unexpected results (i.e. those that were not consistent with their personal or family history), they either dismissed them or they experienced shock or distress. When PRSs were perceived to contradict their health history, some participants cognitively rejected their scores, especially those who trusted their diagnoses. Participant 4 described how they reconciled their low-risk score for autism:

> *“[Impute*.*me calculated a Z score of] like 2*.*5 and they called that medium or low… To me 2*.*5… that’s kind of big, that’s a hell of a risk. 2*.*5, that’s not low… I already had my diagnosis, and it just fits. I like my diagnosis because it helps me explain myself to others*.*”*

Other participants experienced uncertainty, doubt or anxiety when their PRS results did not reflect their personal or family history.

> *“My mother suffers of hypertension but I got a very low score… so is that even possible?” – Participant 2*
>
> *“I have had a lot of anxiety… and I noticed in the polygenic risk score, it was like right in the middle. So I almost thought like, well if I’m as likely as anyone else to get it then, you know, what does that say about me? Is there something wrong with me?” – Participant 11*

Participants who received a high-risk result for a condition that did not run in their family experienced distress – particularly when unexpected results for mental health conditions were received. Participant 6 expressed regretting seeking out a PRS after they received a high-risk score for schizophrenia:

> *“Unfortunately, I do regret getting a PRS… I would have rather not known. I like uncertainty*.*”*

### Behavioural change

Many interviewees reported making behavioural changes based on their PRS results. Importantly, behavioural changes were not limited to those who perceived themselves to be at high risk. Lifestyle changes included weight loss, dietary modifications, more frequent exercise, changes in supplemental or herbal regimen, sunscreen vigilance, practicing self-care, and therapy. Lifestyle changes involving diet, exercise or supplements were implemented as a preventative measure. Therapy and self-care were used as a means of coping with test-related distress stemming from high risk results. Some participants reported feeling more in control of their health after learning about their genetic predisposition to disease, which motivated them to make lifestyle changes.

> *“It made me cautious. But it also made me aware and gave me the power to stop it*.*” – Participant 4*

Another participant described how their PRS results provided direction with what lifestyle changes to make.

> *“It pointed me in a direction where I could develop a game plan and figure out, you know what I was fighting and what was the rational way of approaching it*.*” – Participant 7*

Importantly, not everyone made healthcare changes. One participant felt that their healthy lifestyle left no room for changes, especially in the context of their low-risk results. Another participant made no changes because they did not feel like behavioural changes could offset their genetic risk.

> *“I feel that I can’t do much because there’s something genetic, right? I can’t do much about it*.*” – Participant 2*

## DISCUSSION

To our knowledge, this is the first qualitative study that has engaged with self-initiators of direct-to-consumer polygenic risk scores. We developed a theoretical model that describes participant’s motivations for seeking PRSs, emotional reactions, and actions taken in response to their results. We found that PRS information was often sought as an attempt to compensate for unmet healthcare needs and they accessed this information as a form of self-advoacy. The majority of participants described having a positive experience with receiving PRSs. Polygenic risk scores were often validating for participants, particularly when they aligned with existing personal health concerns or family history. When participants experienced shock and/or distress, it was in relation to the PRS being inconsistent with their expectations. Regardless of their emotional reactions to their results, many participants reported making health-related changes such as dietary modifications, exercise routine, and changes in vitamin supplementation. Elements of our model are supported by the literature, whereas others are more novel findings.

Common motivations for pursuing DTC-GT are general curiosity about genetic information, clarification of disease risk, and identifying opportunities for preventative action(24,28,29). While these motivations were among those described by our study participants, dissatisfaction with healthcare was a core motivator for seeking PRS information. Medical distrust has been previously observed in the context of clinical genetic testing(30), however to our knowledge, unmet healthcare needs have not been noted in relation to DTC-GT. However, DTC-GT research has been largely quantitative, and “dissatisfaction with healthcare” or “unmet healthcare needs” do not appear to have been provided as survey response options. Even when given the option of providing a free text response, selecting “curiosity” may have served as a catch all response for those who couldn’t articulate their motivations. This idea is supported by the data gathered in the context of the pre-interview survey in this study. Although most people had indicated that they were motivated by curiosity, during their interviews they described an underlying medical or emotional reason behind their curiosity. This potential connection between dissatisfaction with healthcare and DTC-GT warrants further exploration.

Our findings suggest that the overall experience with receiving PRSs in a DTC setting is largely positive. Many participants reported feeling validated by their results, particularly when their PRS results aligned with their personal or family history. Participants who received a high-risk score for a pre-existing condition appreciated having an explanation for their diagnoses. While we did not select participants for particular diagnoses, many interview participants were neurodiverse and/or had a history of mental illness. This observation creates an interesting picture when combined with data from previous work showing that psychiatric diagnoses are among the most commonly sought PRSs(7), and data showing that people with these diagnoses are likely to have unmet health care needs(31,32). People who received a high-risk result for a condition that ran in their family were relieved to have confirmation of their genetic risk. When results were not consistent with participant’s personal or family history, many participants found reasons to dismiss or discredit these results. This indicates a role for confirmation bias in responses to self-initated PRS. This is supported by previous work which has identified confirmation bias as relevant reactions to other types of genetic testing(33,34).

Despite describing their experiences as positive, many participants reflected on negative emotions associated with their results. Negative emotions included shock, confusion, anxiety, and concern. The potential for negative reactions contrasts with clinical PRS research that describes mainly positive experiences(16–19,21–23). It is possible that the negative reactions our research showed were mitigated in other studies by the involement of a healthcare provider in pre-test or post-test counseling. Additionally, prior studies provided targeted PRSs to individuals with a personal or family history of cancer(16–21) or bipolar disorder(22). It is possible that having a family history may lead to an overestimation of perceived genetic risk, reducing the burden of a PRS. Rather than receiving targeted PRSs, our participants had a broad range of PRSs available to them, which may explain why more participants reacted negatively to their results. Negative outcomes have also been observed for other DTC third-party interpretation tools(35).

Regardless of participant’s emotional reactions to their results, almost all participants reported making health-related changes in response to their results. As mentioned, behavioural changes were not limited to those who received high-risk results. For many, high-risk results provided motivation to make behavioural changes as an attempt to prevent the onset of disease. Alternatively, low-risk results for pre-existing conditions motivated others to re-evaluate their lifestyle choices. People who did not make lifestyle adjustments were already engaging in healthy behaviours or were not convinced that lifestyle changes could reduce their overall risk. Participants engaged in behavioural changes if they could identify areas in which to improve and when they believed it would make a difference. Our findings contrast with previous PRS research which did not identify behavioural changes made in response to PRS infromation(23,36). It is possible that our study participants were more motivated to make behavioural changes given that their healthcare experiences left them in a state of need.

This study has several limitations. Purposive sampling was used to recruit a demographically diverse group of interviewees. Despite our best efforts, over half of our interview participants were White, in part because the majority Impute.me users are White. Also, because Impute.me users have to overcome the barrier of obtaining and uploading raw genetic information that they received from another source, this population may be considered “early adopters” of this technology and may not represent the average experience with PRS information. Additionally, in accordance with Impute.me’s commitment to preserve anonymity, the researchers did not have access to participant’s PRS results and were unable to evaluate people’s understanding of their results, which could have been useful context for understanding their reactions and responses to their genetic information.

Our research identified a novel relationship between dissatisfaction with healthcare and DTC-GT. More research is needed to explore this finding and moreover, to investigate what underlies “general curiosity” about genetic information. Additionally, as PRSs become incorporated into clinical settings, it will become increasingly important to understand predictors of adverse reactions to PRSs.

We developed a theoretical model describing how unmet healthcare needs and self-advocacy can motivate PRS-seeking behaviour. How people reacted to their results depended on whether their results aligned with their expectations, which were shaped by their lived experience with each condition. Participants used PRSs to inform health-related behavior decisions. The novel relationship between dissatisfaction with healthcare, unmeet need, self-advocacy, and PRSs is important to consider as PRS become integrated into clinical care to maximize the benefit and minimize the harms of PRS information.

## Data Availability

Original qualitative data will not be made publicly available to protect the privacy of research participants.

## CODE AVAILABILITY

Data analyzed through the study is available in the published manuscript. Raw and coded data will not be made available to protect the privacy of research participants

## ACKNOWLEDGEMENTS

The authors offer gratitude to the Coast Salish Peoples, including the x⍰m⍰θkwəy□⍰m (Musqueam), Skwxwú7mesh (Squamish), and Səl□ílw⍰ta⍰/Selilwitulh (Tsleil-Waututh) Nations, on whose traditional, unceded and ancestral territory we have the privilege of working.

## AUTHOR CONTRIBUTIONS

All authors contributed to the design and execution of the study, and the drafting, revision, and final approval of the manuscript. KL, KB, and JA contributed to the analysis and interpretation of the data.

## FUNDING

This research was supported by research grants from the National Society of Genetic Counselors Precision Medicine Special Interest Group and the University of British Columbia and was conducted to fulfill a degree requirement as part of training. JA was supported by the Canada Research Chairs program, and BC Mental Health and Substance Use Services. The funders had no role in review or approval of the manuscript.

## ETHICAL APPROVAL

All procedures followed were in accordance with the ethical standards of the University of British Columbia. Informed consent was obtained from all individual participants included in the study. The study was approved by the University of British Columbia’s Research Ethics Board (H21-01616).

## COMPETING INTERESTS

KL and KB declare no conflicts of interest. Dr. Lasse Folkersen is the founder of Impute.me and Dr. Jehannine Austin provided consultation for the website, though neither made a profit from this service. Dr. Austin also provides consultation for 23andme. The voluntary donations received by Impute.me go to a registered company, from where it is used to pay for server costs. Impute.me was a Danish-law IVS company with ID 37918806, financially audited under Danish tax law.

## Notes

### Author Declarations

All procedures followed were in accordance with the ethical standards of the University of British Columbia. Informed consent was obtained from all individual participants included in the study. The study was approved by the University of British Columbias Research Ethics Board (H21-01616).

