## Supplemental material for "A Qualitative Study Exploring the Consumer Experience of Receiving Self-Initiated Polygenic Risk Scores from a Third-Party Website"

**Interview Guide**

**Can you tell me about what prompted you to seek a PRS from Impute.me?**

- How did you find the Impute.me website?
- Before receiving your results, what did you imagine a polygenic risk score could tell you?
- Was there one genetic condition in particular that you were interested in receiving a PRS for?
- What was your reasoning for your interest in this condition?

*If a current diagnosed condition or something they are at risk for:*

- *Is this something that you have tried to discuss with your doctor?*
- *Do you feel like you’re getting the support/treatment/care from your HCP that you need?*
- *Did you choose to look at your PRS score for this condition because you feel like you’re missing out on something you need for this condition?*
- *Did you look at your PRS scores for this condition as a way to take action?*
- Before receiving the result, what did you think your risk was for this condition?
  - What made you think it was low or high?

**How did you react after receiving your PRS?**

- How did you feel immediately after getting them?
- What did Impute.me estimate as your risk for developing the condition that was of main interest?
  - Was the risk higher or lower than you expected?
- How do the results impact you today? Do you still think about your results today?
- Was there anything you found surprising about your results or how you reacted to them?
- Did you have unanswered questions?
- Who did you tell (friends, family, health care professional, counsellor) about these results?
- Did you find yourself doing anything differently after receiving your PRS? – e.g. what lifestyle changes (diet, exercise, medications, supplements) did you make in response to your results?
  - If lifestyle changes were made, are these changes ongoing?

**Overall, what has your experience of receiving a PRS been like? Was it a positive or negative experience?**

- If positive, would you do it again?
  - Is there anything that you would change about the process that could have made this an even better experience for you?
- If negative, do you regret it?
  - What would you change about the process that could have improved your experience?

**Is there anything else that stood out about your experience that you would like to discuss that I haven’t brought up today?**

*In terms of unanswered questions or emotional support, are there any resources that we can provide you with?*

*We will be analyzing the data from our interviews early next year. Would it be okay if we contact you again if we need anything clarified or have any additional questions for you?*
